# Risk of Myopericarditis following COVID-19 mRNA vaccination in a Large Integrated Health System: A Comparison of Completeness and Timeliness of Two Methods

**DOI:** 10.1101/2021.12.21.21268209

**Authors:** Katie A Sharff, David M Dancoes, Jodi L Longueil, Eric S Johnson, Paul F Lewis

**Affiliations:** Department of Infectious Diseases, Kaiser Permanente Northwest, Portland, Oregon; Department of Analytics, Kaiser Permanente Northwest, Portland, Oregon; Division of Pharmacy, Kaiser Permanente Northwest, Portland, Oregon; Department of Pediatrics, Kaiser Permanente Northwest, Portland Oregon

**Author notes:** Correspondence: Katie A. Sharff, Kaiser Permanente Northwest, Portland, Oregon.

## Abstract

**Purpose:** How completely do hospital discharge diagnoses identify cases of myopericarditis after an mRNA vaccine?

**Methods:** We assembled a cohort 12 to 39 years old patients, insured by Kaiser Permanente Northwest, who received at least one dose of an mRNA vaccine (Pfizer-BioNTech or Moderna) between December 2020 and October 2021. We followed them for up to 30 days after their second dose of an mRNA vaccine to identify encounters for myocarditis, pericarditis or myopericarditis. We compared two identification methods: A method that searched all encounter diagnoses using a brief text description (e.g., ICD-10-CM code I40.9 is defined as ‘acute myocarditis, unspecified’). We searched the text description of all inpatient or outpatient encounter diagnoses (in any position) for “myocarditis” or “pericarditis.” The other method was developed by the Centers for Disease Control and Prevention’s Vaccine Safety Datalink (VSD), which searched for emergency department visits or hospitalizations with a select set of discharge ICD-10-CM diagnosis codes. For both methods, two physicians independently reviewed the identified patient records and classified them as confirmed, probable or not cases using the CDC’s case definition.

**Results:** The encounter methodology identified 14 distinct patients who met the confirmed or probable CDC case definition for acute myocarditis or pericarditis with an onset within 21 days of receipt of COVID-19 vaccination. Three of these 14 patients had an ICD-10 code of I51.4 “Myocarditis, Unspecified” which was overlooked by the VSD algorithm. The VSD methodology identified 11 patients who met the CDC case definition for acute myocarditis or pericarditis. Seven (64%) of the eleven patients had initial care for myopericarditis outside of a KPNW facility and their diagnosis could not be ascertained by the VSD methodology until claims were submitted (median delay of 33 days; range of 12-195 days). Among those who received a second dose of vaccine (n=146,785), we estimated a risk as 95.4 cases of myopericarditis per million second doses administered (95% CI, 52.1 to 160.0).

**Conclusion:** We identified additional valid cases of myopericarditis following an mRNA vaccination that would be missed by the VSD’s search algorithm, which depends on select hospital discharge diagnosis codes. The true incidence of myopericarditis is markedly higher than the incidence reported to US advisory committees. The VSD should validate its search algorithm to improve its sensitivity for myopericarditis.

**Key Points:**

- We identified a higher estimate of myopericarditis following COVID-19 mRNA vaccine by searching encounter text description compared with the Vaccine Safety Datalink (VSD) methodology
- An incomplete list of ICD-10 codes and delays in hospital claims data were responsible for the difference
- We estimated a risk of 95.4 cases of myopericarditis per million second doses administered in patients age 12-39 which is higher than the incidence reported to US advisory committees
- We encourage other VSD sites to validate the case ascertainment of current VSD methodology

## Purpose

Post-marketing vaccine safety in the US is monitored through the complimentary Vaccine Adverse Event Reporting System (VAERS) and the Vaccine Safety Datalink (VSD). VSD has conducted weekly vaccine surveillance known as rapid cycle analysis since the first COVID-19 vaccine was administered in December 2020 to identify rare or serious vaccine related outcomes not identified in clinical trials.(1) This surveillance system looks for serious outcomes associated with 23 pre-specified signals, however the sensitivity of the risk estimates may be limited by only including specific ICD-10 codes, by delays in claims processing when care occurs outside the integrated health system and exclusion of outpatient visits.

Myopericarditis following COVID-19 mRNA vaccination is well reported (2, 3, 4), and the FDA and CDC use VSD analyses of myopericarditis following COVID-19 mRNA vaccination to implement decisions about vaccine policy.

Here we compare the risk of myopericarditis using health record encounter text analysis compared with the VSD rapid cycle analysis methodology from a single integrated health system, Kaiser Permanente Northwest (KPNW), a VSD participant. (1) KPNW represents approximately 7% of the entire VSD population. The purpose of this study was to compare how completely the different methodologies identify post-vaccination myopericarditis.

## Methods

We assembled a cohort of 153,438 adolescents and adults (12 to 39 years old) who were covered by KPNW and were vaccinated with at least one dose of an mRNA vaccine between December 18^th^ 2020 and October 16th 2021. The cohort was followed for up to 30 days after their second dose of an mRNA vaccine to identify encounters for myocarditis, pericarditis or myopericarditis. Encounters included telehealth visits, outpatient visits, including urgent care visits, as well as emergency department visits or hospitalizations. KP’s institutional review board (IRB) approved the study.

KPNW’s electronic health record (EHR) is a version of Epic’s EHR system and captures all encounter diagnoses assigned within KP’s integrated delivery system and affiliated community. The National Center for Health Statistics developed a brief text description or label to define their clinical modification of ICD-10 diagnosis codes. For example, I40.9 is defined as, “acute myocarditis, unspecified”. We searched the text description of all the KPNW encounter diagnoses (in any position) that occurred between December 18^th^ 2020 and October 16, 2021 in both the outpatient and inpatient settings to identify encounters related to “myocarditis” or “pericarditis”. We excluded anyone with a documented diagnosis of myocarditis or pericarditis before their first mRNA vaccination. Two physicians independently reviewed the identified patient records and applied the CDC myocarditis and pericarditis surveillance case definition to classify records as confirmed, probable or not cases based on the prior published definition (5).

To reproduce the VSD methodology (2) we restricted our search to select ICD-10 discharge codes from emergency department visits and hospitalizations, including hospitals owned by KPNW and hospitals unaffiliated with KPNW, which submit insurance claims to KPNW. Each diagnosis associated with a IC10 code of B33.22, B33.23, I30,* or I40* was then flagged as meeting the criteria of being identified by the VSD. As above, two physicians independently reviewed the patient records to classify as confirmed, probable or not cases based on the case definition (5).

We calculated the incidence as a proportion using the person and the dose as the denominator. For brevity, we only present the incidence per million second doses of vaccine. We stratified the incidence by age bands to understand how the risk of myocarditis or pericarditis depended on age. We calculated exact 95% confidence intervals using Stata 17 and the default Clopper-Pearson binomial method (6).

## Results

The encounter text description methodology identified 14 distinct patients ages 12-39 years old who met the confirmed or probable CDC case definition for acute myocarditis or pericarditis within 21 days of receipt of COVID-19 vaccination. Three of these 14 patients had an ICD-10 code of I51.4 Myocarditis, Unspecified which was unique to the encounter text description methodology. When we extended the record look back period to 30 days, we identified 2 additional patients who met the case definition for acute myocarditis or pericarditis within 21 days of COVID-19 vaccination. Although these 2 cases had onset of symptoms within 21 days of vaccination and met the surveillance case definition, the relevant diagnosis code was assigned during an outpatient follow-up visit after the patient was discharged from the emergency department or hospital.

Using the VSD methodology, we identified 11 patients ages 12-39 years old who met the CDC case definition for acute myocarditis or pericarditis within 21 days of receipt of COVID-19 vaccination. Seven (64%) of the eleven patients had initial care for myopericarditis outside of a KPNW facility and their diagnosis could not be ascertained by the VSD methodology until claims were submitted. Four of the eleven cases identified by the VSD had claims data submitted after 30 days, with an average claims delay of 64 days and a median delay of 33 days (range 12-195 days). Claims data for these events were submitted from patient encounters at community hospitals that are not owned by Kaiser Permanente Northwest but provide care for our patients. (Table 1)

**Table 1:** Summary of Myopericarditis cases Identified by Encounter Text Description and VSD Methodology

| Patient | VSD method | Reason Missed | Days to Claim Filed | Age | Sex | Vaccine | Dose | Days to chest pain onset | EKG | Trop peak mcg/L | Evaluation of CAD | LVEF on echo, % or cardiac MRI | LOS, d |
| --- | --- | --- | --- | --- | --- | --- | --- | --- | --- | --- | --- | --- | --- |
| 1 | No | I51.4 not queried |  | 18-24 | M | Pfizer | 2 | 3 | Sinus arrhythmia | 4.08 | Not done | 57%-MRI | 1 |
| 2 | No | I51.4 not queried |  | 18-24 | M | Pfizer | 2 | 4 | Sinus | 2.9 | Not done | 70% | 2 |
| 3 | No | I51.4 not queried |  | 18-24 | M | Pfizer | 2 | 1 | Sinus Tach | 11.1 | Not done | 40% | 2 |
| 4 | No | Short lookback |  | 12-17 | M | Pfizer | 2 | 4 | Sinus | .04 | Not done | Normal | ED |
| 5* | No | Short lookback |  | 30-39 | F | Pfizer | 1 | 1 | Antero-lateral ischemia | <=.03 | Normal angio | 60-65% | OP |
| 6 | Yes, > 30 days | Delay in claim | 33 | 12-17 | M | Pfizer | 2 | 2 | ST elevation | 4.02 | Not done | 62%-MRI | 2 |
| 7 | Yes, > 30 days | Delay in claim | 74 | 12-17 | M | Pfizer | 2 | 3 | ST elevation | 12.4 | Not done | 61%-MRI | 5 |
| 8* | Yes, > 30 days | Delay in claim | 82 | 30-39 | M | Pfizer | 2 | 15 | ST elevation | <=.03 | Not done | Not done | ED |
| 9* | Yes, > 30 days | Delay in claim | 195 | 12-17 | M | Pfizer | 1 | 7 | ST abnormality | 0.5 | Normal angio | 22% | 43 |
| 10 | Yes |  | Internal^ | 12-17 | M | Pfizer | 2 | 3 | Sinus | 13.3 | Not done | Normal | 3 |
| 11 | Yes |  | 29 | 12-17 | M | Pfizer | 2 | 3 | ST elevation | 66.13 | Not done | 27%-MRI | 4 |
| 12 | Yes |  | 22 | 18-24 | M | Pfizer | 2 | 3 | Sinus | 0.38-Trop T | Not done | 55-60% | ED |
| 13 | Yes |  | Internal^ | 18-24 | F | Moderna | 2 | 4 | ST elevation, sinus tach | 15.9 | Not done | 60% | 1 |
| 14 | Yes |  | Internal^ | 18-24 | M | Pfizer | 2 | 3 | Sinus | 5.19 | Not done | 68%-MRI | 1 |
| 15 | Yes |  | 12 | 18-24 | M | Pfizer | 2 | 3 | ST elevation | 4.9 | Not done | 50% | 1 |
| 16 | Yes |  | Internal^ | 18-24 | M | Moderna | 2 | 3 | ST elevation, lateral | 9.62 | Not done | 48-55% | 1 |

<sup>^</sup>Internal: Internal hospital encounter, claims data not used to identify case
LVEF: Left-ventricular end-systolic function
OP: outpatient
ED: Emergency Department
Normal troponin Range: <=.03 mcg/L
\*Cases that met case definition but were atypical from general pattern

In our patients ages 12-39 years old who received a second dose of vaccine (n=146,785), we estimated a risk of 95.4 cases of myopericarditis per million second doses administered (95% CI, 52.1 to 160.0). In males who received a second dose (n=66,533) we estimated a rate of 195.4 cases of myopericarditis per million second doses (95% CI, 104.0 to 334.1). (Figure 1)

**Figure 1:**
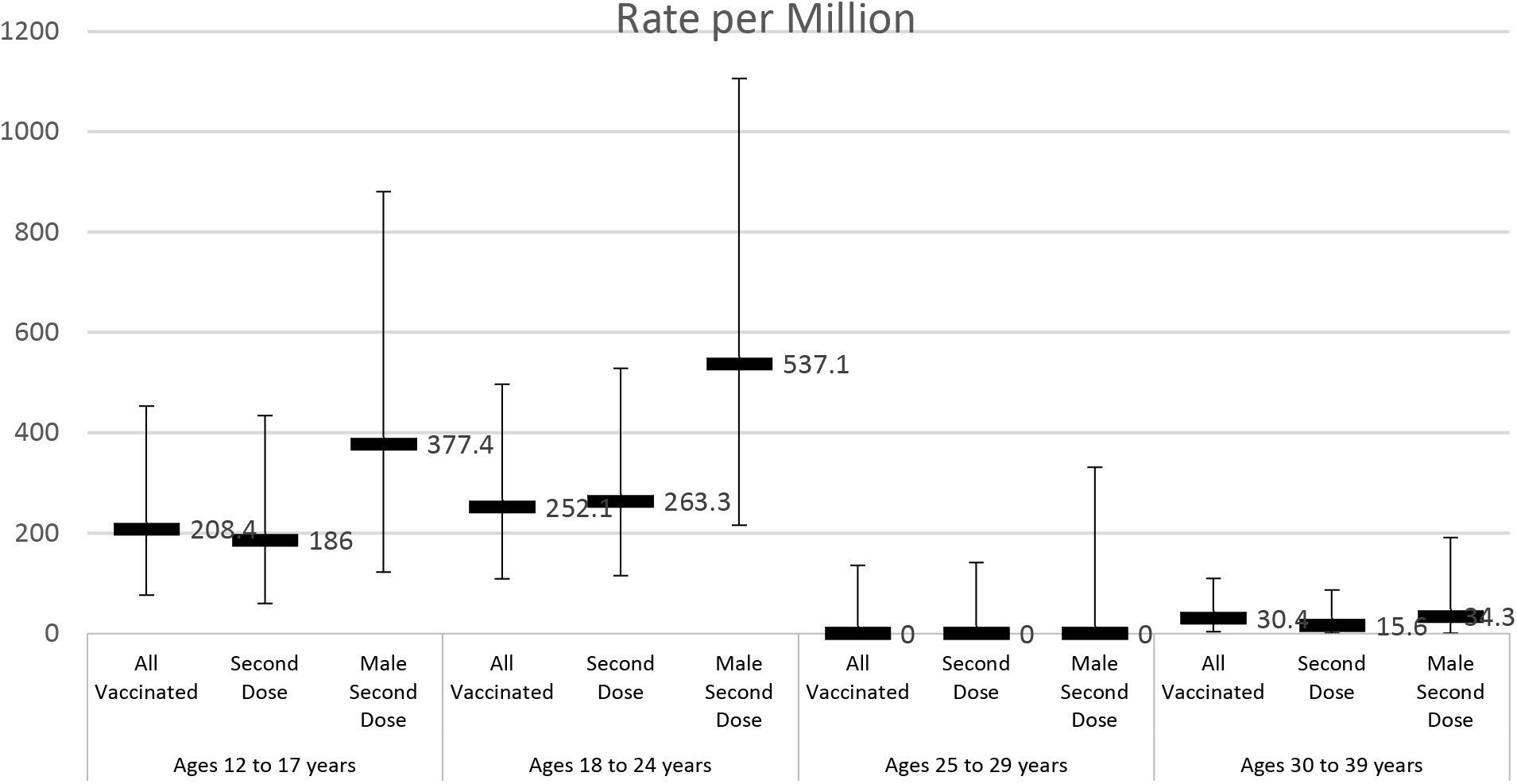
Rate per Million

## Conclusion

We identified a higher estimate of myopericarditis following COVID-19 mRNA vaccine by searching encounter text description in the medical record of an integrated health system compared with the VSD methodology. The VSD specifically excluded ICD-10 code I51.4, Myocarditis, Unspecified resulting in missed episodes that met the case definition. Additionally, some of the VSD sites rely on claims data from community hospitals that are not owned by the health systems to identify cases. Although their methodology may eventually identify these cases if the analysis were conducted several months after all the events occurred, the lag in claims submission and payment may result in inaccurate case estimates. Finally, by extending our look back period with the encounter text description method to 30 days after the episode, we were able to capture additional events that met case definition but were not coded in the health record by day 21. For example, one patient had a hospital discharge code of Chest Pain (ICD-10 code R07.9), that occurred 4 days after vaccination, however when he had follow-up in the pediatric cardiology clinic the appropriate diagnosis code of Idiopathic Myocarditis (ICD-10 code I40.1) was documented. This outpatient pediatric cardiology appointment occurred at day 25 after his initial presentation.

Our estimate of the incidence of myopericarditis following COVID-19 mRNA vaccine is similar in magnitude to that reported from two studies from Israel (7, 8) but higher than that reported in the US studies and at VBRPAC and ACIP meetings (4, 9, 10, 11). Complete case estimates are essential when modeling risk and benefit for wide-scale vaccine implementation and booster doses in younger age groups.

Using an identical population, we identified that the encounter text description methodology identified approximately twice as many cases of myopericarditis following COVID-19 mRNA vaccination. The VSD is a multi-site consortium with several sites relying on outside claims data to identify cases, potentially resulting in prolonged data lags for accurate ascertainment of events. We would encourage other VSD sites to validate the case ascertainment of current VSD methodology. Future modeling and public policy decisions on vaccine safety should consider the sensitivity limitations of VSD derived estimates.

## Data Availability

All data produced in the present study are available upon reasonable request to the authors

## References

1. Klein NP, Lewis N, Goddard K, et al. Surveillance for Adverse Events after COVID-19 mRNA Vaccination. JAMA. 2021 Oct 12;326(14):1390–1399.

2. Marshall M, Ferguson ID, Lewis P, Jaggi P, Gagliardo C, Collins JS, Shaughnessy R, Caron R, Fuss C, Corbin KJE, Emuren L, Faherty E, Hall EK, Di Pentima C, Oster ME, Paintsil E, Siddiqui S, Timchak DM, Guzman-Cottrill JA. Symptomatic Acute Myocarditis in 7 Adolescents After Pfizer-BioNTech COVID-19 Vaccination. Pediatrics. 2021 Sep;148(3):e2021052478.

3. Kim HW, Jenista ER, Wendell DC, Azevedo CF, Campbell MJ, Darty SN, Parker MA, Kim RJ. Patients With Acute Myocarditis Following mRNA COVID-19 Vaccination. JAMA Cardiol. 2021 Oct 1;6(10):1196–1201.

4. Montgomery J, Ryan M, Engler R, Hoffman D, McClenathan B, Collins L, Loran D, Hrncir D, Herring K, Platzer M, Adams N, Sanou A, Cooper LT Jr. Myocarditis Following Immunization With mRNA COVID-19 Vaccines in Members of the US Military. JAMA Cardiol. 2021 Oct 1;6(10):1202–1206.

5. Gargano JW, Wallace M, Hadler SC, Langley G, Su JR, Oster ME, Broder KR, Gee J, Weintraub E, Shimabukuro T, Scobie HM, Moulia D, Markowitz LE, Wharton M, McNally VV, Romero JR, Talbot HK, Lee GM, Daley MF, Oliver SE. Use of mRNA COVID-19 Vaccine After Reports of Myocarditis Among Vaccine Recipients: Update from the Advisory Committee on Immunization Practices -United States, June 2021. MMWR Morb Mortal Wkly Rep. 2021 Jul 9;70(27):977–982.

6. Clopper, C. J., Pearson ES. The use of confidence or fiducial limits illustrated in the case of the binomial. Biometrika 1934; 26:404–413.

7. Mevorach D, Anis E, Cedar N, Bromberg M, Haas EJ, Nadir E, Olsha-Castell S, Arad D, Hasin T, Levi N, Asleh R, Amir O, Meir K, Cohen D, Dichtiar R, Novick D, Hershkovitz Y, Dagan R, Leitersdorf I, Ben-Ami R, Miskin I, Saliba W, Muhsen K, Levi Y, Green MS, Keinan-Boker L, Alroy-Preis S. Myocarditis after BNT162b2 mRNA Vaccine against Covid-19 in Israel. N Engl J Med. 2021 Dec 2;385(23):2140–2149.

8. Witberg G, Barda N, Hoss S, Richter I, Wiessman M, Aviv Y, Grinberg T, Auster O, Dagan N, Balicer RD, Kornowski R. Myocarditis after Covid-19 Vaccination in a Large Health Care Organization. N Engl J Med. 2021 Dec 2;385(23):2132–2139.

9. Simone A, Herald J, Chen A, Gulati N, Shen AY, Lewin B, Lee MS. Acute Myocarditis Following COVID-19 mRNA Vaccination in Adults Aged 18 Years or Older. JAMA Intern Med. 2021 Dec 1;181(12):1668–1670.

10. Matthew Oster, Sara Oliver, Proceedings of the Advisory Committee on Immunization Practices, 11/2/2021. ACIP November 2-3, 2021 Presentation Slides | Immunization Practices | CDC)

11. Matthew Oster, Proceedings of the Vaccines and Related Biological Products Advisory Committee, 10/26/21. Vaccines and Related Biological Products Advisory Committee October 26, 2021 Meeting Announcement -10/26/2021 - 10/26/2021 | FDA

